# Sativex (Nabiximols) for the treatment of Agitation & Aggression in Alzheimer’s Dementia in UK nursing homes (STAND): a randomised, double-blind, placebo-controlled feasibility trial

**DOI:** 10.1101/2024.12.18.24319032

**Authors:** Christopher P Albertyn, Ta-Wei Guu, Petrina Chu, Byron Creese, Allan H Young, Latha Velayudhan, Sagnik Bhattacharyya, Hassan Jafari, Simrat Kaur, Pooja Kandangwa, Ben Carter, Dag Aarsland

## Abstract

**Background:** Alzheimer’s Disease (AD) patients often experience clinically significant agitation, leading to distress, increased healthcare costs, and earlier institutionalisation. Current treatments have limited efficacy and significant side effects. Cannabinoid-based therapies, such as the nabiximols oral spray (brand name: Sativex®; 1:1 delta-9-tetrahydrocannabinol (THC) and cannabidiol (CBD)), offer potential alternatives.

**Methods:** The ‘Sativex® for Agitation & Aggression in Alzheimer’s Dementia’ (STAND) trial was a randomised, double-blind, placebo-controlled, feasibility study conducted in UK care homes. Participants with probable AD and significant agitation were randomised to receive placebo or nabiximols for 4 weeks on an up-titrated schedule, followed by a 4-week observation period. This trial is registered with ISRCTN 7163562.

**Findings:** Between October 2021 and June 2022, 53 candidates were assessed; 29 met eligibility criteria and were randomised. No participants withdrew, and adherence was high, and was generally feasible to deliver. The intervention was well tolerated, with no safety concerns reported.

**Interpretation:** Despite significant COVID-19 pandemic related challenges, administering nabiximols to advanced AD patients with agitation demonstrated feasibility and safety. Although no statistically significant treatment effects were observed, indications of positive clinical effects were noted. These findings support further investigation into cannabinoid-based therapies for agitation in AD.

## Introduction

Alzheimer’s disease (AD), a progressive neurodegenerative disorder, is characterised by a decline in cognitive function, which significantly impacts daily living and quality of life [1]. Among the myriad of symptoms associated with AD, behavioural and psychological symptoms of dementia (BPSD) are particularly challenging. BPSD encompasses a range of non-cognitive disturbances including agitation, aggression, delusions, hallucinations, depression, and nocturnal sleep disturbances. These symptoms are prevalent in up to 90% of individuals with dementia at some point during their illness [2]. Agitation, a common manifestation of BPSD, is particularly distressing for both patients and caregivers. It is characterised by excessive motor activity, verbal aggression, and physical aggression, often leading to increased caregiver burden, higher healthcare costs, and accelerated institutionalization [3,4].

In the UK, care home residents with dementia frequently exhibit agitation, posing significant challenges to care providers. Approximately 70% of care home residents in the UK have dementia, with a substantial proportion experiencing severe BPSD [5]. The management of agitation in this population is complex, often requiring a combination of pharmacological and non-pharmacological interventions. However, current pharmacological treatments, including most antipsychotics and benzodiazepines, are associated with limited efficacy and significant adverse effects, such as increased risk of falls, sedation, and cardiovascular events [6,7]. Consequently, there is an urgent need for safer and more effective therapeutic options.

Recent advances in cannabinoid psychopharmacology have highlighted the potential of cannabinoid-based therapies in managing BPSD, particularly agitation [8]. Cannabinoids, the active compounds found in the cannabis plant, interact with the endocannabinoid system, which plays a crucial role in regulating mood, cognition, and behaviour [9–11]. Nabiximols (Sativex®), an oromucosal spray containing a balanced ratio of delta-9-tetrahydrocannabinol (THC) and cannabidiol (CBD), has emerged as a promising candidate for this purpose. It is currently licensed for the treatment of spasticity in multiple sclerosis, but its potential benefits may extend beyond this indication [12,13].

In this study, we aimed to explore the feasibility of nabiximols for people living with AD and agitation in care homes.

## Methods

### Study design

The STAND study was a double-blind, parallel-group, randomised placebo-controlled trial assessing the feasibility of nabiximols (Sativex) for treating agitation and aggression in Alzheimer’s disease dementia. The trial received ethics approval from West Midlands – Coventry & Warwickshire Research Ethics Committee). The protocol, Statistical Analysis Plan and study materials are available [14,15].

### Participants

Participants aged 55 to 95 with a probable AD diagnosis, and clinically significant agitation/aggression (A/A), were recruited from existing care homes within the greater London area, primarily through the NIHR Maudsley Biomedical Research Centre Care Home Research Network (CHRN). Full eligibility criteria can be sourced from the protocol [14] and ISRCTN preregistration [16]. Witnessed informed consent was obtained from the participant, or a personal/professional legal representative either in-person, via post, or via electronic consent.

### Randomisation and masking

At the end of baseline assessments, participants were randomised (1:1) to receive either nabiximols sprays containing 2·7mg THC / 2·5mg CBD or placebo sprays with the same peppermint oil flavouring/colourings. Randomisation was stratified by baseline AD severity (low: Functional Assessment Staging (FAST) ≤5, moderate: FAST=6, severe: FAST=7) and the sequence was generated using randomly varying block (sizes of two and four) by King’s Clinical Trials Unit (KCTU) and hosted on a web-based system hosted by KCTU. Treatment allocation was blinded to study researchers, participants, family caregivers, and care home staff. Care home staff nurses administered sprays according to a standardised up-titrating dosing schedule.

All parties remained blinded to the treatment allocation until after the data lock and presentation of the study results. The trial statistician was kept blinded until approval of the Statistical Analysis Plan (SAP) by the independent Trial Steering Committee (TSC), at which point the trial statistician was able to review data partially blinded (arms labelled A/B) to prepare the analysis code. After database lock, the trial statistician was able to request data fully unblinded to prepare the final report.

### Procedures

The target dose was 4 sprays/day of nabiximols (10.8mg THC/10mg CBD) or placebo, titrated up from 1 spray per day for the first 3 days, to a maximum dose of 4 sprays per day. The intervention was up-titrated for a total treatment duration of 4 weeks, with participants receiving 1 spray in the morning, 1 spray in the afternoon, and 2 sprays in the evening for the latter 2 weeks. The participants were then checked 2 and 4 weeks post-treatment for safety and other outcome measures. The researchers completed outcome assessments concerning safety, adherence and clinical outcomes at baseline, weeks 2, 4, 6 and 8 either virtually or in-person. Participants with issues such as side-effects, physical conditions, or non-compliance limiting dose-escalation, were checked by the study doctor and the principal investigator to confirm whether they should remain on the current dose or be withdrawn from the trial.

### Outcomes

The primary objective was to assess feasibility, defined by four key criteria: randomising 60 participants within 12 months, achieving a ≥75% follow-up rate at 4 weeks, maintaining ≥80% adherence to allocation, and demonstrating a clinically meaningful effect size (≥0.3) on the Cohen-Mansfield Agitation Inventory (CMAI) score.

Safety and tolerability were assessed through self- and carer-reported side effects, adverse events, and suicidality (Columbia-Suicide Severity Rating Scale, C-SSRS). Secondary clinical outcomes included CMAI and Neuropsychiatry Inventory-Nursing Home Version (NPI-NH) scores, collected fortnightly from weeks 0-8. Additional secondary outcomes, such as Quality of Life (QOL-AD, QUALID), Abbey Pain Scale (APS), standardised Mini Mental State Examination (sMMSE), were collected at baseline, week 4, and week 8. Functional Assessment Staging of Alzheimer’s Disease (FAST), and Clinical Frailty Scale (CFS) scores were collected at baseline and week 4 only.

### Sample size and Statistical analysis

A sample size of 60 (1:1) would have enabled study researchers to estimate a drop-out rate of 20% to within a 95% confidence interval of ±10%. Primary feasibility analyses were conducted on all available data.

The trial steering committee took approved the statistical analysis plan prior to the analyst gaining access to the trial arm (coded A/B) following KCTU SOPs. A modified intention to treat analysis was used for clinical outcomes that included randomised participants with a baseline and at least one follow-up measurement of the outcome. Analysis was conducted on Stata version 18.

For the primary feasibility outcomes, the number and relevant proportions of participants randomised, retained, and deemed adherent were reported alongside 95% confidence intervals (CIs) specifying exact binomial distributions.

Demographics and secondary outcome measures were summarised using descriptive statistics by arm and overall at each timepoint. The mean difference between arms for secondary outcome measures were estimated using mixed linear models adjusting for the fixed effects of arm, baseline disease severity, baseline measurement of the outcome, time, and an arm*time interaction term. Marginal treatment effects were reported for each follow-up timepoint with an adjusted mean difference (aMD). A random intercept was fitted at the participant level to account of repeated measurements. For secondary outcomes only measured at baseline and 1 follow-up timepoint (i.e. FAST and CFS), no fixed effect for time and arm*time was included and no random intercept was included. For heavily skewed continuous measures, the relevant score categories were used instead in similar logistic regression models. Standardised treatment effect estimates (Cohen’s d) for continuous outcomes were calculated by dividing mean differences between arms by the pooled baseline standard deviation (SD) of the outcome. No significance level was set and no p-values were reported as the aim was to provide a range of preliminary effect estimates.

## Results

Between 13^th^ October 2021 and 30^th^ June 2022, 53 nursing home residents consented and were assessed for eligibility. The COVID-19 pandemic, particularly the Omicron wave, disrupted recruitment, leading to reduced screening and a pause in trial activities. Ultimately, 29 participants (55% of those assessed) were randomised after excluding 24 ineligible participants (14 to placebo, 15 to nabiximols). All randomised participants completed the 8-week follow-up period and were included in primary analyses (Figure 1).

**Figure 1.**
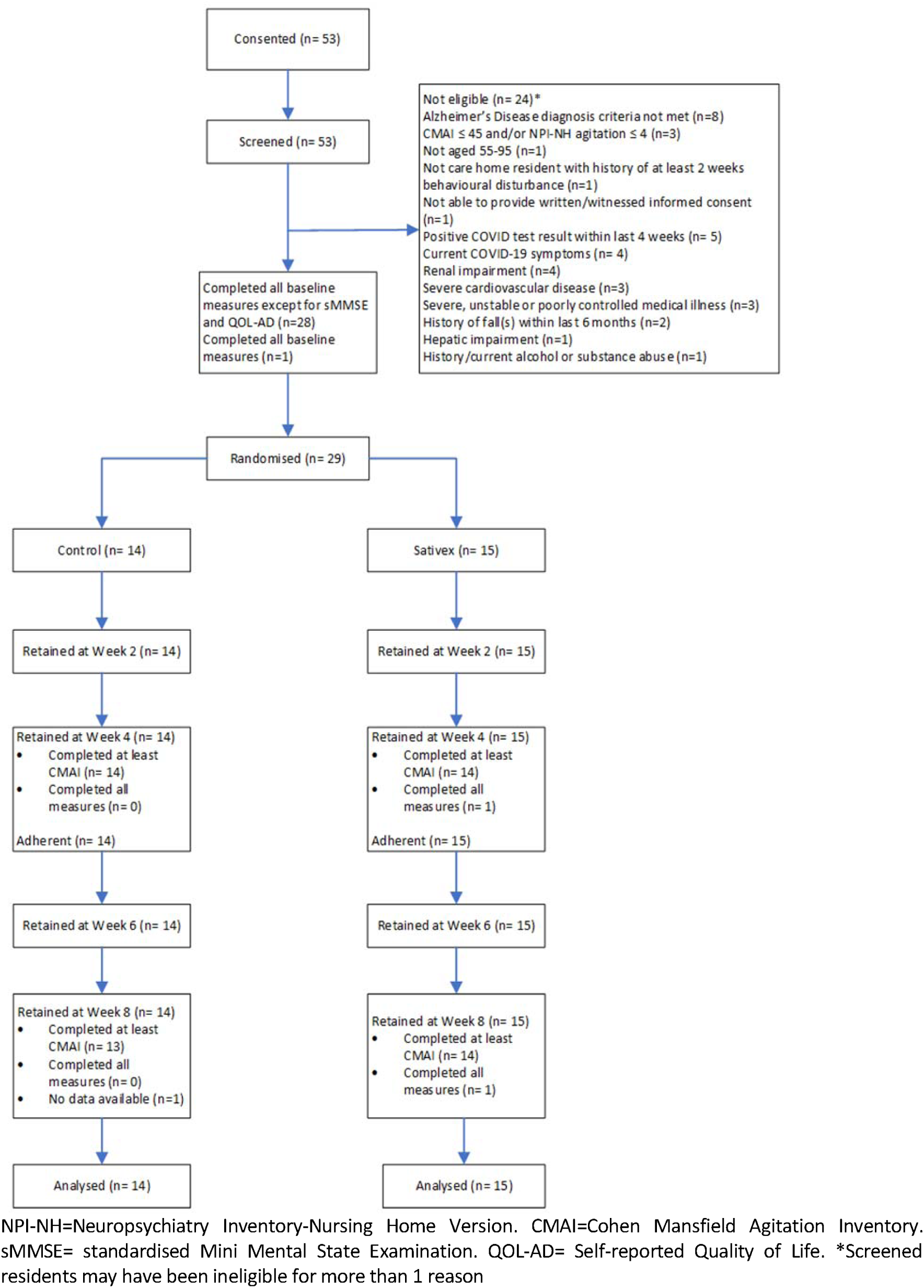
CONSORT diagram depicting the participant recruitment journey and randomisation allocation.

Table 1 shows baseline demographic and clinical characteristics of the participants. The two groups were similar at baseline in terms of age, sex, severity of cognitive decline and Abbey pain scale, but the nabiximols group had higher baseline agitation severity and overall BPSD scores.

**Table 1.** Baseline demographics and clinical characteristics of participants.

| Demographics/characteristics |  | Nabiximols (n= 15) | Placebo (n= 14) |
| --- | --- | --- | --- |
| Age* |  | 81.7 (7.9) | 80 (8.6) |
| Sex |  |  |  |
|  | Male | 6 (40%) | 9 (64%) |
|  | Female | 9 (60%) | 5 (36%) |
| Disease severity |  |  |  |
|  | Moderate (FAST ≤ 5) | 0 (0%) | 0 (0%) |
|  | Moderately Severe (FAST = 6) | 5 (33%) | 4 (29%) |
|  | Severe (FAST = 7) | 10 (67%) | 10 (71%) |
| Mean CMAI total score (SD) |  | 95.5 (29.0) | 71.5 (26.3) |
| Mean NPI-NH total score (SD) |  | 58.5 (24.3) | 30.2 (20.9) |
| Mean NPI-NH disruptiveness score (SD) |  | 16.3 (11.4) | 10.2 (8.4) |
| APS category |  |  |  |
|  | No pain | 14 (93%) | 12 (86%) |
|  | Mild pain | 1 (7%) | 2 (14%) |
|  | Moderate pain | 0 (0%) | 0 (0%) |
|  | Severe pain | 0 (0%) | 0 (0%) |

Data are n (%). \*The age at randomisation was skewed, and the median (IQR) of the two groups were 83.0 (77.0-86.0) and 81.0 (75.0-87.0), respectively. FAST=Functional Assessment Staging Tool for Dementia. CMAI=Cohen Mansfield Agitation Inventory. NPI-NH=Neuropsychiatry Inventory-Nursing Home Version. APS=Abbey Pain Scale.

The primary feasibility outcome measures are summarised in Table 2. Retention was 100% (29/29) at both Week 4 and Week 8 follow-ups, surpassing the 75% target. Adherence was 100% (29/29), participants received at least one dose per week and generally adhered to the titration schedule (see Supplementary Table 1 for the titration progress and summaries of intervention experiences). However, the clinical effect size for CMAI did not reach the desired threshold (Cohen’s d ≥ -0·3), showing 0·23 at Week 4 and 0·08 at Week 8.

**Table 2.**
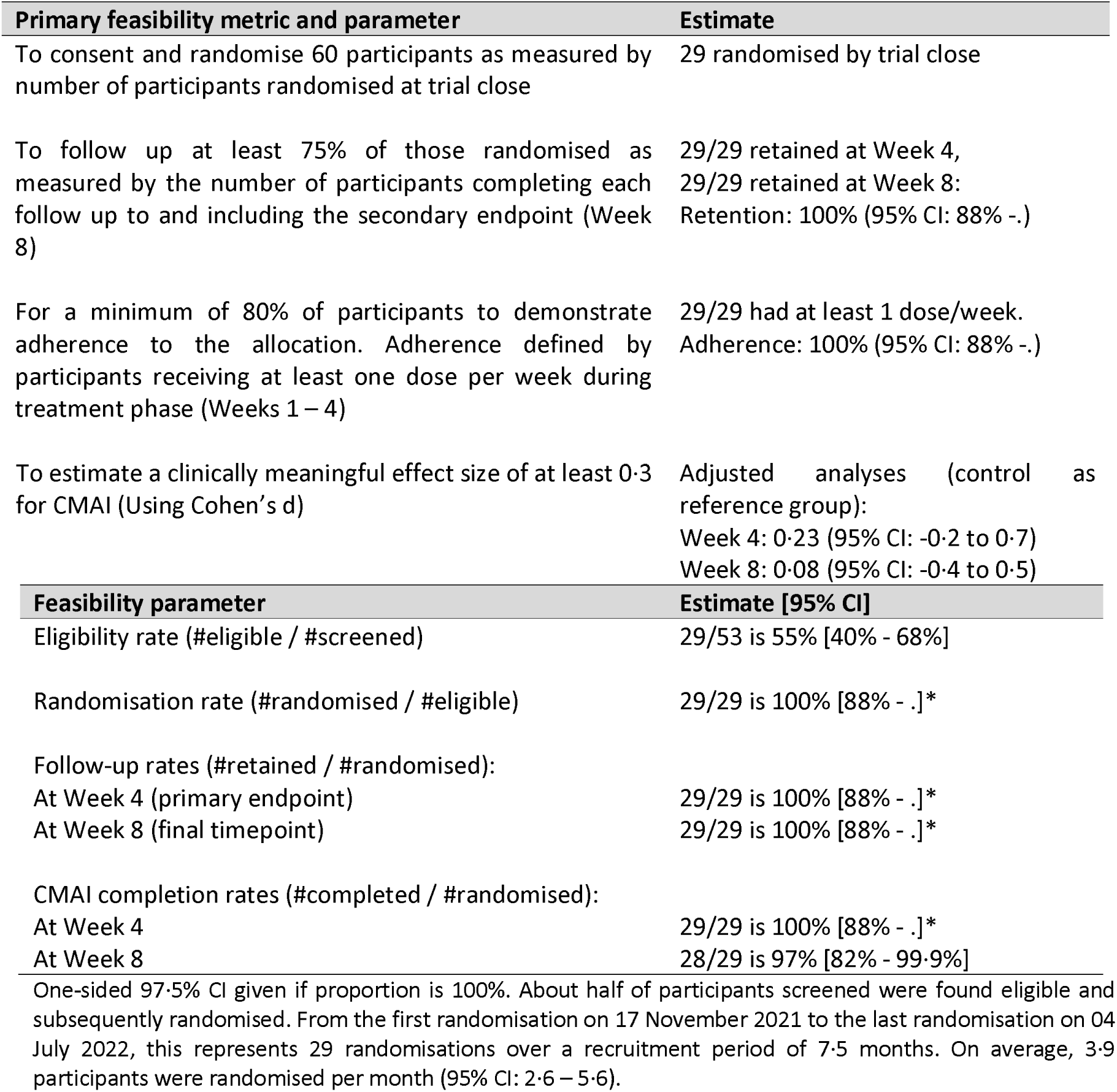
STAND primary feasibility progression outcome measures.

| Primary feasibility metric and parameter | Estimate |
| --- | --- |
| To consent and randomise 60 participants as measured by number of participants randomised at trial close | 29 randomised by trial close |
| To follow up at least 75% of those randomised as measured by the number of participants completing each follow up to and including the secondary endpoint (Week 8) | 29/29 retained at Week 4,<br>29/29 retained at Week 8:<br>Retention: 100% (95% CI: 88% -.) |
| For a minimum of 80% of participants to demonstrate adherence to the allocation. Adherence defined by participants receiving at least one dose per week during treatment phase (Weeks 1 – 4) | 29/29 had at least 1 dose/week.<br>Adherence: 100% (95% CI: 88% -.) |
| To estimate a clinically meaningful effect size of at least 0.3 for CMAI (Using Cohen's d) | Adjusted analyses (control as reference group):<br>Week 4: 0.23 (95% CI: -0.2 to 0.7)<br>Week 8: 0.08 (95% CI: -0.4 to 0.5) |
| Feasibility parameter | Estimate [95% CI] |
| Eligibility rate (#eligible / #screened) | 29/53 is 55% [40% - 68%] |
| Randomisation rate (#randomised / #eligible) | 29/29 is 100% [88% - .]* |
| Follow-up rates (#retained / #randomised): |  |
| At Week 4 (primary endpoint) | 29/29 is 100% [88% - .]* |
| At Week 8 (final timepoint) | 29/29 is 100% [88% - .]* |
| CMAI completion rates (#completed / #randomised): |  |
| At Week 4 | 29/29 is 100% [88% - .]* |
| At Week 8 | 28/29 is 97% [82% - 99.9%] |
| One-sided 97.5% CI given if proportion is 100%. About half of participants screened were found eligible and subsequently randomised. From the first randomisation on 17 November 2021 to the last randomisation on 04 July 2022, this represents 29 randomisations over a recruitment period of 7.5 months. On average, 3.9 participants were randomised per month (95% CI: 2.6 – 5.6). |  |

In this trial, 21% (6 of 29) of participants reported at least one non-serious adverse event (AE), all were mild severity, and unrelated to the study intervention. The AE listing included hepatobiliary, respiratory, eye, and skin/tissue disorders, with mild cases across the placebo (n=2) and nabiximols (n=4) arms. Serious adverse events (SAEs) occurred in 2 participants (1 in nabiximols arm, and 1 participant in placebo arm with 2 events), with moderate or severe respiratory and renal/urinary disorders. No AEs nor SAEs were attributed to the intervention, and there were no reported deaths, falls, increase in C-SSRS or withdrawals (see supplementary Table 2 for summary of adverse events and serious adverse events by trial arm).

The results of the adjusted analysis for secondary clinical and neuropsychiatric outcomes are shown in Table 3, and the full secondary neuropsychiatric outcome data by trial arm and timepoint can be found in the Supplementary materials (Supplementary Table 3 and Supplementary Figures). At the end of the treatment period by week 4, CMAI scores had improved in both groups, with mean scores reducing to 58·2 (SD 23·9) in the control and 77·0 (SD 24·5) in the nabiximols group. There was no association between nabiximols and total CMAI at Week 4 with an adjusted mean difference (aMD) of 6·77 (95% CI -6·71 to 20·25; Cohen’s d=0·23), or four weeks post-treatment at Week 8. Similar trends were observed in NPI-NH scores, which decreased in both groups at week 4 and further by week 8, as shown in Figure 2. QUALID and FAST scores showed no significant changes between groups, and the adjusted effect sizes did not reach clinically meaningful thresholds. By week 8, mean adjusted differences between nabiximols and control groups for CMAI, NPI-NH, and QUALID remained minimal (CMAI: 2·43; NPI-NH: 2·53; QUALID: -2·61).

**Figure 2.**
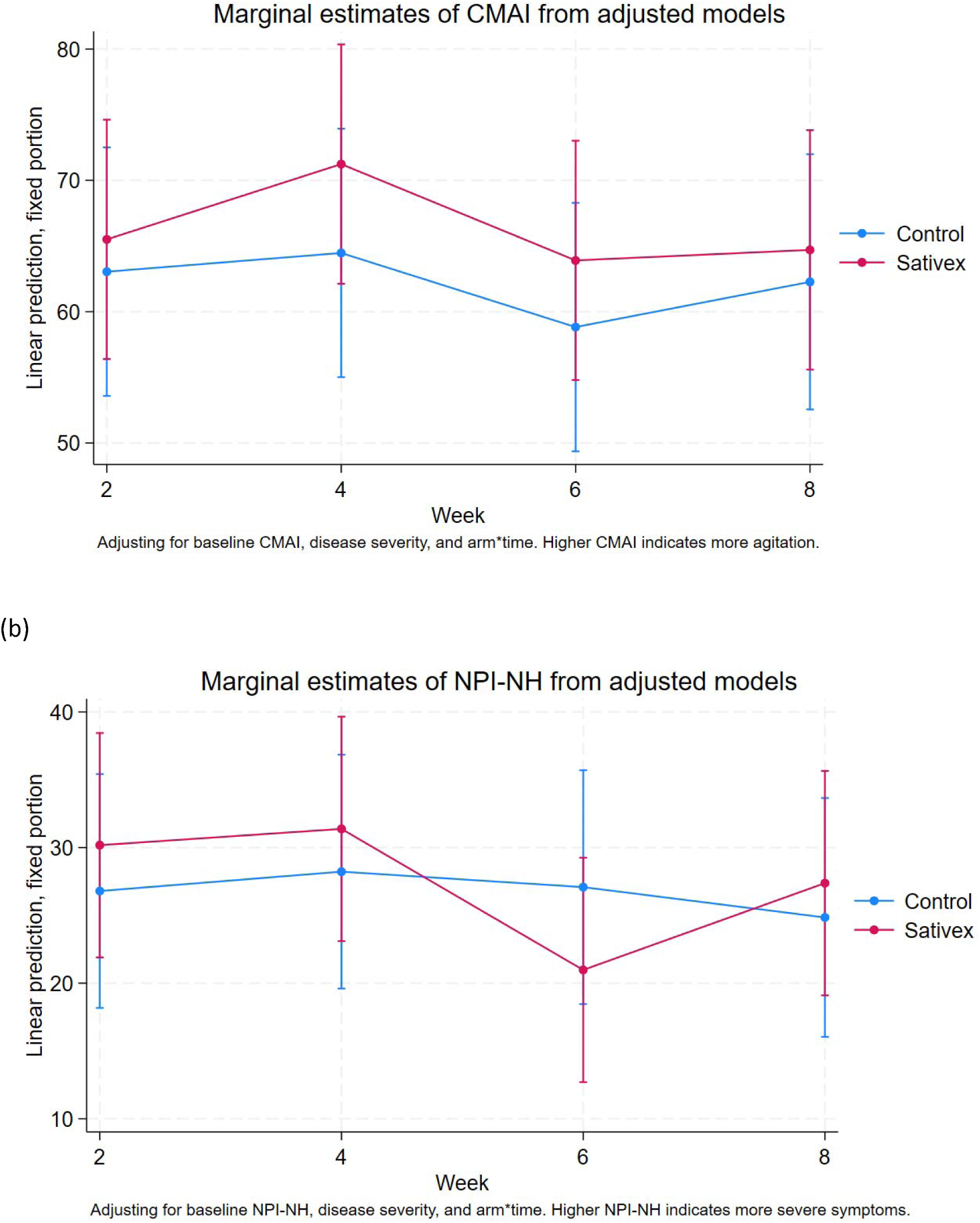
Marginal estimate for CMAI (a) and NPI-NH (b) scores by treatment arm.

**Table 3.** Results of adjusted analyses for secondary clinical and neuropsychiatric outcomes.

| **Outcome** | **N in analysis**<br>**model** | **Mean difference (95% CI)** | **Cohen *d***<br>**(95% CI)** |
| --- | --- | --- | --- |
| **CMAI** | 29 |  |  |
| **4 weeks** |  | 6.77 (-6.71 , 20.25) | 0.23 (-0.23 , 0.68) |
| **8 weeks** |  | 2.43 (-11.23 , 16.10) | 0.08 (-0.38 , 0.54) |
| **NPI-NH** | 29 |  |  |
| **4 weeks** |  | 3.15 (-9.46 , 15.76) | 0.12 (-0.36 , 0.59) |
| **8 weeks** |  | 2.53 (-10.2 , 15.26) | 0.10 (-0.38 , 0.57) |
| **QOL-AD** | 0 |  |  |
| **4 weeks** |  | N/A | N/A |
| **8 weeks** |  | N/A | N/A |
| **QUALID** | 29 |  |  |
| **4 weeks** |  | -0.25 (-4.21 , 3.7) | -0.03 (-0.47 , 0.42) |
| **8 weeks** |  | -2.61 (-6.63 , 1.41) | -0.29 (-0.75 , 0.16) |
| **APS\*** | 29 |  |  |
| **4 weeks** |  | 1.08 (0 , .)** | N/A |
| **8 weeks** |  | 3.82 (0.28 , 52.09)** | N/A |
| **sMMSE** |  |  |  |
| **4 weeks** |  | N/A | N/A |
| **8 weeks** |  | N/A | N/A |
| **FAST at 4 weeks** | 29 | -1.22 (-2.23 , -0.21) | -0.71 (-1.30 , -0.12) |
| **CFS at 4 weeks\*** | 19 | 1.13 (0.06 , 21.09)*** | N/A |

\*Odds ratios (ORs) reported instead of mean differences for APS and CFS measures.
\*\*All participants had reported APS scores in the 'No pain' category at Week 4, there were no values in the 'Mild pain' cells. There were only 4 participants reporting APS scores in the 'Mild pain' category at Week 8.
\*\*\*Only 2 participants reported CFS scores in either the 'Living with very mild frailty' or 'Mild frailty' categories at Week 4 and some observations were omitted because they perfectly predicted being in the combined lower categories.

Pain levels, as measured by the APS, remained low across groups, with no moderate or severe pain reported throughout the study. A small decrease (improvement) in dementia function staging was observed at Week 4 (aMD: -1·22 [-2·23, -0·21]) but only two participants in the nabiximols arm actually improved enough at Week 4 to move from the ‘severe’ disease category to the ‘moderately severe’ disease category.

## Discussion

The STAND trial demonstrated the feasibility of conducting a randomized, placebo-controlled study of cannabinoid oral spray for agitation in patients with Alzheimer’s disease. Despite under-recruitment, most screened patients participated, with high retention and adherence throughout the trial. No adverse events attributable to the intervention were identified, and the safety profile appeared to be similar between the nabiximols arm and the placebo arm.

The recruitment challenges should be considered in the context of recruiting in care homes during the COVID pandemic. Critically, the initial planned 12-month recruitment window was reduced to 9-months, resulting in 3 fewer months than originally planned to reach the recruitment target of 60 participants. Even within these 9-months, recruitment had been paused because of the Omicron wave in the earlier months. Accounting for only having 75% of the planned recruitment window, intermittent trial pauses, and with our recruitment rate accelerating in the later months, it may be that the sample target would have been reached if the trial was operational for the full planned 12-months. The unprecedented burnout among care home staff globally limited the capacity for clinical research[17], even as residents likely experienced escalating BPSD [18]. Additionally, the pandemic contributed to attrition through increased mortality, participant ineligibility due to presence of covid symptoms, and care home closures across the UK, all of which impacted the recruitment and randomisation of the study. Despite challenges, retention reached 100%, and data completion for key neuropsychiatric outcomes was robust, supporting the feasibility of administering this intervention.

With regard to the safety profile, although all the participants in this study were classified as either moderately severe or severe dementia, and the majority of the participants (100% in placebo arm and 73% in nabiximols arm) had moderate to severe frailty, there were very few reports of adverse events. A similar study conducted by our team, applying pure CBD of a much higher dosage (up to 600mg daily) in a modest sample, found that 12.5% of participants in the CBD arm had treatment-related dizziness [19]. Another small-sized study done in Australia using an oral spray of higher dosage THC/CBD mixture (up to 50lllmg THC, 34lllmg CBD daily), also found a significantly higher rate of overall adverse events in their treatment arm [20]. Compared to these studies, ours applying low dose THC/CBD appeared to be safer and better tolerated, even in a cohort of frail, late-stage AD patients.

Safety remains the foremost concern in designing interventions for BPSD. Antipsychotics, benzodiazepines, and antidepressants are commonly used empirically for these symptoms. However, prior to the approval of brexpiprazole—a partial agonist at dopamine D2 and D3 receptors—for agitation in Alzheimer’s disease, no medication was considered safe for long-term use [21]. Even brexpiprazole carries a black box warning, highlighting the increased risk of mortality associated with antipsychotic use in elderly patients with dementia-related psychosis [7]. Benzodiazepines are known for their negative impact on the cognition and postural balance of the elderly [22]. Antidepressants like citalopram and mirtazapine have limitations too: citalopram may reduce agitation but can impair cognition and prolong QT interval [23], while mirtazapine showed no benefit for agitation in a large RCT and might increase mortality risk [24]. Given the favourable safety and tolerability profile shown at the current dose, we would recommend a further dose-finding study to establish the minimal effective dose before initiating a larger phase-III trial. The STAND trial focused on a nursing home population, allowing for continuous safety and compliance monitoring. However, based on our positive safety results, future studies should consider including community-dwelling participants to enhance recruitment and reach a broader patient population in need.

While the adjusted analyses indicated little evidence of a signal from nabiximols, this study was not powered to demonstrate a clinical effect. The small sample size and lack of precision should be considered when interpreting the effectiveness of the intervention, along with the finding that only half of participants completed the full titration schedule as planned. In addition, agitation is likely a heterogeneous and multi-factorial syndrome that needs to be defined and measured with more specificity, as recommended previously by the SYMBAD study [24]. Although using the questionnaires such as the NPI and the CMAI for agitation assessment have been widely validated and implemented, wearable device has been proposed to complement these traditional instruments [25,26]. These device-based measurements might have the potential to reveal information associated with the outcome not easily captured by the questionnaires and therefore stratify the patients and outcomes more specifically. For example, the International Psychogeriatric Association recently suggested the recognition and separation of nocturnal/circadian agitation [3], wearable devices capable of continuous behavioural measurement might have the potential to distinguish symptoms happen in daytime and at night, and therefore facilitate the outcome measurement.

This trial introduces two key innovations relevant to the feasibility, safety and efficacy to be considered for future studies: the use of an oromucosal spray for drug delivery and the application of cannabinoids in dementia care. It has been noted in a target product profile developed by a consortium of experts that an ideal therapeutic for agitation in AD would address difficulty swallowing (dysphagia) and refusal to swallow [27]. Our study is the largest to examine the oro-mucosal administration of medication for agitation in people living with AD, addressing common challenges like dysphagia and refusal to swallow, which likely contributed to the high tolerability and adherence observed. In addition, future studies might explore nabiximols as an adjunct to sensory interventions, such as music, light and aroma therapy [28,29], leveraging cannabis’s potential to enhance sensory experiences and regulate circadian rhythm [30,31].^29,30^ This combined cannabinoid-sensory approach could enable gradual deprescription, supporting long-term, non-pharmacological management of agitation in AD.

The primary limitations of this study include under-recruitment due to the COVID-19 pandemic, which reduced the statistical power needed to determine effect size. Although our sample was slightly larger than those in comparable studies and featured a more balanced sex distribution, the participant numbers remain insufficient to robustly estimate the intervention’s effect size. Additionally, while the relatively low doses of THC and CBD likely contributed to fewer adverse events compared with prior studies, these dosages may also be less effective in reducing agitation and aggressive behaviours. The limited sample size warrants cautious interpretation of the safety and efficacy profile in the nabiximols group. Finally, the ethnic homogeneity of our sample may limit the generalisability of these findings beyond the UK and Ireland, particularly given the high proportion of white British participants.

This study has three main strengths: a robust design, favourable safety profile, and high retention rate. Strict inclusion and exclusion criteria, while reducing participant numbers, safeguarded this vulnerable population from serious adverse events and minimized infection risk among participants, caregivers, and researchers. The observed safety profile in this frail, late-stage dementia cohort likely contributed to the 100% retention rate, underscoring the intervention’s feasibility even amid pandemic-related pressures on care home staff.

In conclusion, this study demonstrates the feasibility of a pilot randomized, placebo-controlled trial of nabiximols oral spray for agitation in Alzheimer’s disease patients in care homes, with no safety concerns observed. The findings provide insights that can inform future trials in refining dosing schedules and minimizing adverse event risks. However, establishing the intervention’s effect size will require further investigation to assess therapeutic efficacy more definitively.

## Data sharing statement

A de-identified version of the dataset for meta-analyses or analyses will be available with investigator support from 1 year after the publication upon reasonable request to the authors. Written proposals will be assessed by the Principal Investigator, Co-Investigators, and members of the Kings Clinical Trials Unit (KCTU) committee, and a decision about the appropriateness of the use of data will be made. A data sharing agreement would need to be put in place before any data is shared.

## Data Availability

A de-identified version of the dataset for meta-analyses or analyses will be available with
investigator support from 1 year after the publication upon reasonable request to the authors. Written proposals will be assessed by the Principal Investigator, Co-Investigators, and members of the Kings Clinical Trials Unit (KCTU) committee,
and a decision about the appropriateness of the use of data will be made. A data sharing
agreement would need to be put in place before any data is shared.

## Acknowledgements

This research is funded by Alzheimer’s Research UK (Registered Charity No: 207711 (England & Wales), SC041156 (Scotland)), Global Clinical Trials Fund (GCTF). Additional support is supplied from the Maudsley NIHR Biomedical Research Centre (BRC), the NIHR Clinical Research Network (CRN), and Ageing Research at King’s (ARK; www.kcl.ac.uk/ark). The IMP and placebo were provided by Jazz Pharmaceuticals (previously GW pharmaceuticals). The funders had no role in study design, data collection and analysis, decision to publish, or preparation of the manuscript. The views expressed are those of the authors and not necessarily those of the NIHR or the Department of Health and Social Care.

We would also like to acknowledge the significant contribution of Dr Iskandar Johar, who was instrumental in the conception, development and operation of the STAND trial, but sadly passed before this publication was drafted.

## CRediT Taxonomy of Authors’ contributions

CRediT Taxonomy of Authors' contributions
| Contribution | Author |
| --- | --- |
| Conceptualisation | CPA, B Creese, AY, LV, SB, B Carter, DA [IJ] |
| Data Curation | PC, HJ, B Carter |
| Formal analysis | PC, B Carter |
| Funding acquisition | CPA, AY, LV, SB, B Carter, DA |
| Investigation | CPA, SK, PK, TWG |
| Methodology | CPA, B Creese, AY, LV, SB, B Carter, DA [IJ] |
| Project administration | CPA, AY, LV, SB, B Carter, DA |
| Software | n/a |
| Resources | CPA |
| Supervision | CPA, B Creese, AY, LV, SB, B Carter, DA [IJ] |
| Validation | AY, LV, SB, B Carter, DA |
| Visualization | PC, B Carter |
| Writing – original draft | CPA, TWG, PC |
| Writing – review & editing | B Creese, AY, LV, SB, B Carter, DA |

