## Supplementary materials for "Sativex (Nabiximols) for the treatment of Agitation & Aggression in Alzheimer’s Dementia in UK nursing homes (STAND): a randomised, double-blind, placebo-controlled feasibility trial"

### Appendix

**Supplementary Information: Questionnaires and information used for patient selection and outcome assessment**

**Study materials, SPIRIT checklist and full Statistical Analysis Plan:** https://doi.org/10.6084/m9.figshare.23260934.v1

**Diagnosis of Alzheimer's Disease:** McKhann G, Drachman D, Folstein M, Katzman R, Price D, Stadlan EM. Clinical diagnosis of Alzheimer's disease: report of the NINCDS-ADRDA Work Group under the auspices of Department of Health and Human Services Task Force on Alzheimer's Disease. *Neurology* 1984; **34**(7): 939-44.

**CMAI:** Cohen-Mansfield J. Assessment of agitation. *Int Psychogeriatr* 1996; **8**(2): 233-45.

**NPI-NH:** Cummings J. The Neuropsychiatric Inventory: Development and Applications. *J Geriatr Psychiatry Neurol* 2020; **33**(2): 73-84.

**FAST:** Auer S, Reisberg B. The GDS/FAST staging system. *Int Psychogeriatr* 1997; **9 Suppl 1**: 167-71.

**C-SSRS:** Posner K, Brown GK, Stanley B, et al. The Columbia-Suicide Severity Rating Scale: initial validity and internal consistency findings from three multisite studies with adolescents and adults. *Am J Psychiatry* 2011; **168**(12): 1266-77.

**QoL-AD:** Beer C, Flicker L, Horner B, et al. Factors associated with self and informant ratings of the quality of life of people with dementia living in care facilities: a cross sectional study. *PloS one* 2010; **5**(12): e15621.

**QUALID:** Bond J. Quality of life for people with dementia: approaches to the challenge of measurement. *Ageing and Society* 1999; **19**(5): 561-79.

**APS:** Abbey J, Piller N, Bellis AD, et al. The Abbey pain scale: a 1-minute numerical indicator for people with end-stage dementia. *International journal of palliative nursing* 2004; **10**(1): 6-13.

**sMMSE:** Tombaugh TN, McIntyre NJ. The mini-mental state examination: a comprehensive review. *J Am Geriatr Soc* 1992; **40**(9): 922-35.

**CFS:** Rockwood K, Abeysundera MJ, Mitnitski A. How should we grade frailty in nursing home patients? *J Am Med Dir Assoc* 2007; **8**(9): 595-603.

**Supplementary Table 1: Summaries of intervention experiences by trial arm and overall**

| Intervention experience | Control (N= 14) | Sativex (N= 15) | Overall (N= 29) |
|---|---|---|---|
| Number completing titration schedule as planned (%) [95% CI] | 6 (43%) [18 – 71%] | 10 (67%) [38 – 88%] | 16 (55%) [36 – 74%] |
| **Week 1 (12 total)** | | | |
| Missed doses | 5 (36%) | 4 (27%) | 9 (31%) |
| Took total scheduled doses | 8 (57%) | 11 (73%) | 19 (66%) |
| Took more doses than scheduled | 1 (7%) | 0 (0%) | 1 (3%) |
| **Week 2 (21 total)** | | | |
| Missed doses | 4 (29%) | 3 (20%) | 7 (24%) |
| Took total scheduled doses | 10 (71%) | 12 (80%) | 22 (76%) |
| **Week 3 (28 total)** | | | |
| Missed doses | 3 (21%) | 5 (33%) | 8 (28%) |
| Took total scheduled doses | 11 (79%) | 10 (67%) | 21 (72%) |
| **Week 4 (28 total)** | | | |
| Missed doses | 3 (21%) | 4 (27%) | 7 (24%) |
| Took total scheduled doses | 11 (79%) | 11 (73%) | 22 (76%) |

One-sided 97.5% CI given if proportion is 100%. Although all participants were defined as adherent, just over half of all randomised participants completed the titration schedule as planned. There was one participant in the control arm that was accidentally given additional evening doses during the first two days.

**Supplementary Table 2: Summary of adverse events and serious adverse events by trial arm**

| Adverse event | Control (N= 2) | Sativex (N= 4) | Overall (N= 6) |
|---|---|---|---|
| Category | | | |
| Hepatobiliary disorders | 1 | 2 | 3 |
| Respiratory disorders | 0 | 1 | 1 |
| Eye disorders | 1 | 0 | 1 |
| Skin/tissues disorders | 0 | 1 | 1 |
| Severity | | | |
| 1. Mild | 2 (100%) | 4 (100%) | 6 (100%) |
| Relationship to study intervention | | | |
| 4. Remote | 0 (0%) | 1 (25%) | 1 (17%) |
| 5. Not related | 2 (100%) | 3 (75%) | 5 (83%) |
| **Serious adverse event** | **Control (N= 2)** | **Sativex (N= 1)** | **Overall (N= 3)** |
| Category | | | |
| Respiratory, thoracic, and mediastinal disorders | 1 | 0 | 1 |
| Renal and urinary disorders | 0 | 1 | 1 |
| Unknown category | 1 | 0 | 1 |
| Severity | | | |
| 2. Moderate | 0 (0%) | 1 (100%) | 1 (33%) |
| 3. Severe | 2 (100%) | 0 (0%) | 2 (67%) |
| Relationship to study intervention | | | |
| 5. Not related | 2 (100%) | 1 (100%) | 3 (100%) |

If a single number is presented, the number of events and number of unique reporters are the same. In this case, all events were reported by different participants. No events were deemed to be related to study intervention, and there were no falls, withdrawals or deaths were reported. The event description for the 1 SAE of unknown system organ class was ‘Atrial Fibrillation’. The two SAEs in the control arm were reported for the same participant.

**Supplementary Table 3***: Secondary neuropsychiatric outcome data by trial arm and timepoint*

| | BASELINE | | | WEEK 4 (PRIMARY ENDPOINT) | | | WEEK 8 (FINAL TIMEPOINT) | | |
|---|---|---|---|---|---|---|---|---|---|
| **Outcome** | **Control (N=14)** | **Sativex (N=15)** | **Overall (N=29)** | **Control (N=14)** | **Sativex (N=15)** | **Overall (N=29)** | **Control (N=14)** | **Sativex (N=15)** | **Overall (N=29)** |
| **CMAI total – median (IQR)** *[missing (% of N)]* | 65.0 (53.0-78.0) | 84.0 (78.0-109.0) | 78.0 (63.0-106.0) | 49.5 (45.0-61.0) | 81.0 (57.0-93.0) | 61.0 (48.0-83.0) | 44.0 (43.0-64.0) *[1 (7%)]* | 62.0 (49.0-94.0) | 59.5 (43.5-75.5) *[1 (3%)]* |
| **CMAI total – mean (SD)** *[missing (% of N)]* | 71.5 (26.3) | 95.5 (29.0) | 83.9 (29.8) | 58.2 (23.9) | 77.0 (24.5) | 67.9 (25.6) | 55.5 (21.4) *[1 (7%)]* | 70.5 (24.2) | 63.6 (23.8) *[1 (3%)]* |
| **NPI-NH total – mean (SD)** *[missing (% of N)]* | 30.2 (20.9) | 58.5 (24.3) | 44.8 (26.6) | 22.5 (16.7) | 36.6 (18.2) | 29.8 (18.6) | 18.7 (15.0) *[1 (7%)]* | 32.6 (20.7) | 26.1 (19.3) *[1 (3%)]* |
| **NPI-NH disruptiveness – mean (SD)** *[missing (% of N)]* | 10.2 (8.4) | 16.3 (11.4) | 13.4 (10.4) | 7.3 (5.5) | 13.3 (6.2) | 10.4 (6.5) | 6.8 (4.4) *[1 (7%)]* | 11.5 (7.5) | 9.3 (6.6) *[1 (3%)]* |
| **QoL-AD total – mean (SD)** *[missing (% of N)]* | . (.) *[14 (100%)]* | 43.0 (.) *[14 (93%)]* | 43.0 (.) *[28 (97%)]* | 15.0 (.) *[13 (93%)]* | 36.0 (1.4) *[13 (87%)]* | 29.0 (12.2) *[26 (90%)]* | 23.0 (.) *[13 (93%)]* | 38.0 (4.2) *[13 (87%)]* | 33.0 (9.2) *[26 (90%)]* |
| **QUALID total – mean (SD)** *[missing (% of N)]* | 29.9 (9.4) | 33.3 (8.4) | 31.6 (8.9) | 27.4 (9.6) | 29.8 (8.9) | 28.6 (9.2) | 28.5 (10.6) *[1 (7%)]* | 28.4 (7.5) | 28.5 (8.9) *[1 (3%)]* |
| **APS category – n (%)** | | | | | | | | | |
| **No pain** | 12 (86%) | 14 (93%) | 26 (90%) | 14 (100%) | 15 (100%) | 29 (100%) | 12 (86%) | 12 (80%) | 24 (83%) |
| **Mild pain** | 2 (14%) | 1 (7%) | 3 (10%) | 0 (0%) | 0 (0%) | 0 (0%) | 1 (7%) | 3 (20%) | 4 (14%) |
| **Moderate pain** | 0 (0%) | 0 (0%) | 0 (0%) | 0 (0%) | 0 (0%) | 0 (0%) | 0 (0%) | 0 (0%) | 0 (0%) |
| **Severe pain** | 0 (0%) | 0 (0%) | 0 (0%) | 0 (0%) | 0 (0%) | 0 (0%) | 0 (0%) | 0 (0%) | 0 (0%) |
| *[missing (% of N)]* | | | | | | | *[1 (7%)]* | | *[1 (3%)]* |

| | BASELINE | | | WEEK 4 (PRIMARY ENDPOINT) | | | WEEK 8 (FINAL TIMEPOINT) | | |
|---|---|---|---|---|---|---|---|---|---|
| **Outcome** | **Control (N=14)** | **Sativex (N=15)** | **Overall (N=29)** | **Control (N=14)** | **Sativex (N=15)** | **Overall (N=29)** | **Control (N=14)** | **Sativex (N=15)** | **Overall (N=29)** |
| **sMMSE total – mean (SD)** *[missing (% of N)]* | . (.) *[14 (100%)]* | 17.0 (.) *[14 (93%)]* | 17.0 (.) *[28 (97%)]* | . (.) *[14 (100%)]* | 18.0 (.) *[14 (93%)]* | 18.0 (.) *[28 (97%)]* | . (.) *[14 (100%)]* | 16.0 (.) *[14 (93%)]* | 16.0 (.) *[28 (97%)]* |
| **FAST (stratifier categories) - n (%)** | | | | | | | | | |
| **Moderately Severe** | 4 (29%) | 5 (33%) | 9 (31%) | 4 (29%) | 7 (47%) | 11 (38%) | | | |
| **Severe** | 10 (71%) | 10 (67%) | 20 (69%) | 10 (71%) | 8 (53%) | 18 (62%) | | | |
| **CFS summary category – n (%)** | | | | | | | | | |
| **Living with very mild frailty** | 0 (0%) | 0 (0%) | 0 (0%) | 0 (0%) | 1 (7%) | 1 (3%) | | | |
| **Mild frailty** | 0 (0%) | 4 (27%) | 4 (14%) | 1 (7%) | 0 (0%) | 1 (3%) | | | |
| **Moderate/severe frailty** | 14 (100%) | 11 (73%) | 25 (86%) | 13 (93%) | 14 (93%) | 27 (93%) | | | |

The CMAI, NPI-NH, FAST, CFS, APS, and QUALID measures were completed at all timepoints, except where one participant in the control arm did not have available responses for any measures collected at Week 8.

Almost no responses were collected for the QOL-AD or sMMSE at any timepoint due to multiple refusals from care home residents to conduct the assessments. At baseline, only 1 participant provided responses to the QOL-AD and sMMSE. At Weeks 4 and 8, only 3 participants had scored responses for the QOL-AD. At Weeks 4 and 8, there was only 1 participant that had scored responses for the sMMSE. If a future trial is planned in the same population, it may be inadvisable to include the QOL-AD and sMMSE as study measures.

There may be some baseline imbalance between arms for the total CMAI and NPI-NH scores. No significance testing was carried out to formally assess baseline imbalance, only visual inspection of boxplots.

All participants reported APS scores falling in the ‘No pain’ or ‘Mild pain’ categories at each timepoint.

There were 4 participants in Sativex that reported worsened CFS scores at Week 4, all 4 of these participants had high enough scores to be categorised in a higher frailty category (from ‘Mild frailty’ at baseline to ‘Moderate/severe frailty’ at Week 4). One participant in the Sativex arm reported improved frailty, moving from the ‘Moderate/severe frailty’ category at baseline to the ‘Living with very mild frailty’ category at Week 4. One participant in the control arm reported improved frailty, moving from the ‘Moderate/severe frailty’ category at baseline to the ‘Mild frailty’ category at Week 4.

**Supplementary Figure 1 and 2. The crude CMAI and NPI-NH scores in the trial period.**

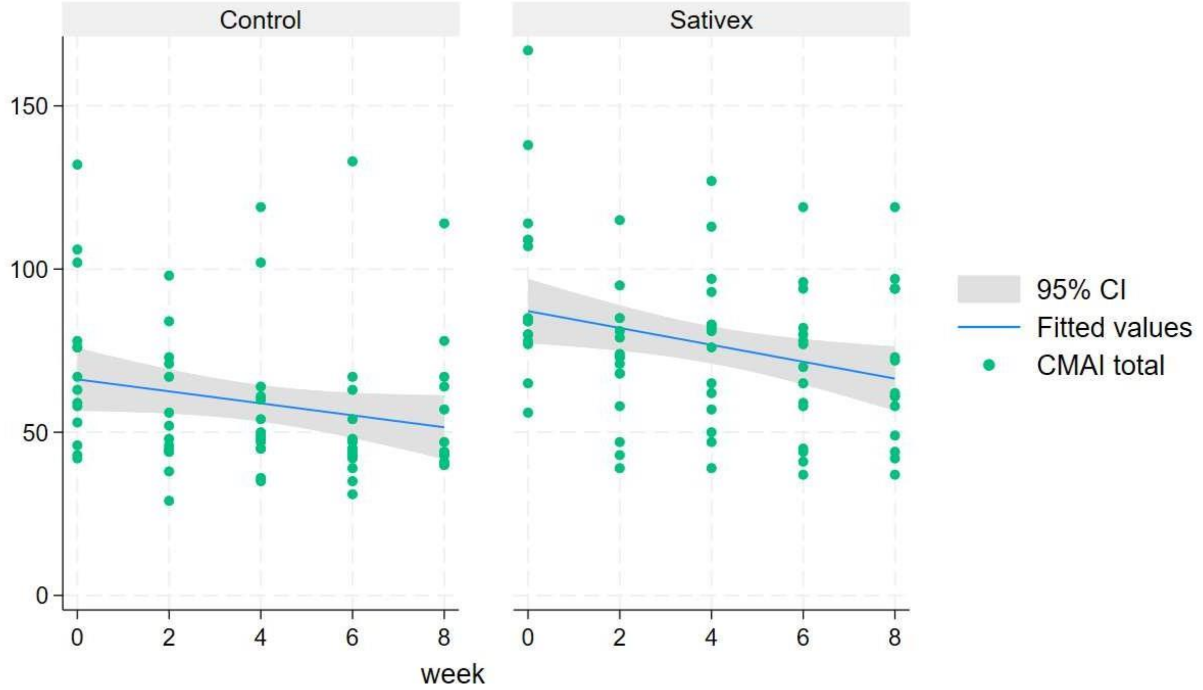


Supplementary Figure 1. CMAI totals by arm over 8 weeks of follow-up

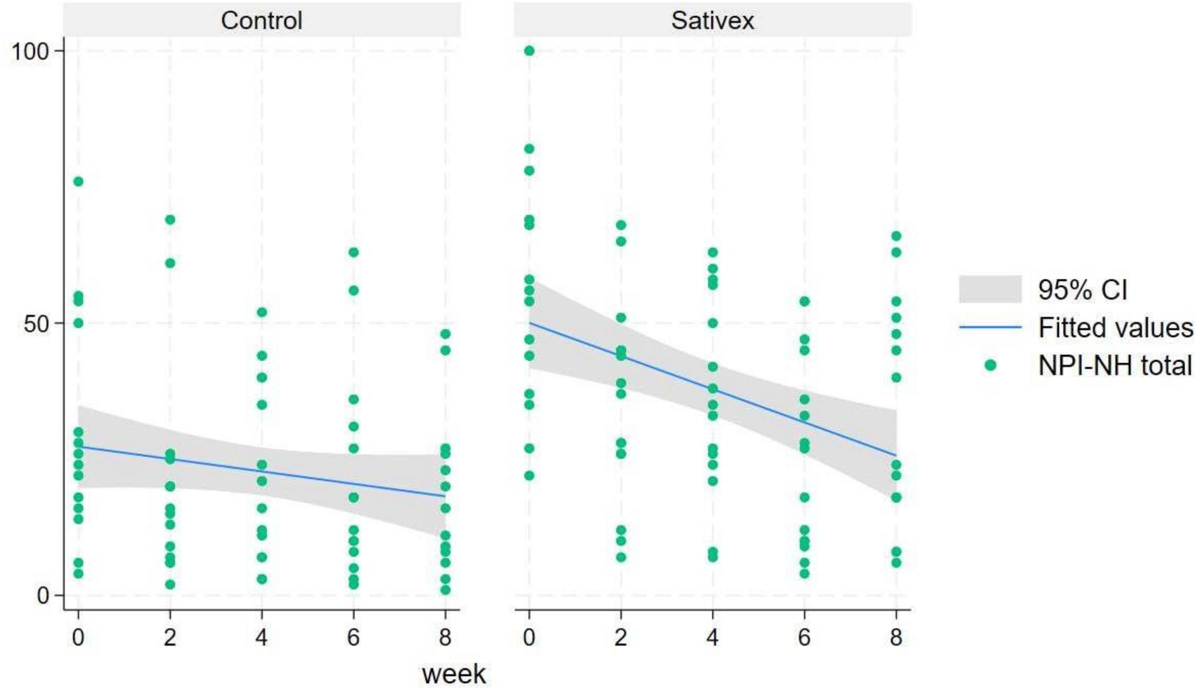


Supplementary Figure 2. NPI-NH totals by arm over 8 weeks of follow-up

The crude CMAI and NPI-NH total scores assessed during the 8-week trial period. Supplementary Figures 1 and 2 show the total scores (crude) over the 8 weeks of follow-up by trial arm. Both the crude CMAI and NPI-NH total scores appeared higher at baseline in the Satives arm.
